# Probabilistic Graphical Models for Evaluating the Utility of Data-Driven ICD Code Categories in Pediatric Sepsis

**DOI:** 10.1101/2024.10.01.24314724

**Authors:** Lourdes A. Valdez, Edgar Javier Hernandez, O’Connor Matthews, Matthew A. Mulvey, Hillary Crandall, Karen Eilbeck

**Affiliations:** University of Utah, Salt Lake City, UT

## Abstract

Electronic health records (EHRs) are digitalized medical charts and the standard method of clinical data collection. They have emerged as valuable sources of data for outcomes research, offering vast repositories of patient information for analysis. Definitions for pediatric sepsis diagnosis are ambiguous, resulting in delayed diagnosis and treatment, highlighting the need for precise and efficient patient categorizing techniques. Nevertheless, the use of EHRs in research poses challenges. EHRs, although originally created to document patient encounters, are now primarily used to satisfy billing requirements. As a result, EHR data may lack granularity, potentially leading to misclassification and incomplete representation of patient conditions. We compared data-driven ICD code categories to chart review using probabilistic graphical models (PGMs) due to their ability to handle uncertainty and incorporate prior knowledge. Overall, this paper demonstrates the potential of using PGMs to address these challenges and improve the analysis of ICD codes for sepsis outcomes research.

## Introduction

EHRs are the digitalized medical charts and the standard method of clinical data collection. They contain valuable information such as patient medical history, demographics, diagnosis, treatments, and procedures, including imaging and lab results. They are potentially a powerful resource for research because they are cost-effective and enable rapid collection of large data sets for retrospective studies and trend observation in most disease spaces (1–3). EHR data, primarily the diagnostic and procedure codes, have been used in many studies, including outcomes research, risk analysis, disease prediction, drug efficacy and use, and healthcare resource utilization and medical costs (4–24).

The International Classification of Diseases (ICD) is an established set of diagnosis codes used to document and bill patient encounters, facilitating record-keeping between healthcare providers and clinics (25). ICD codes are widely used in research as proxies for clinical outcomes. However, their administrative nature makes this a cause for concern among many healthcare professionals. Differences in coding practices across institutions, discrepancies between EHR documentation behavior and the real world, and the financial motivations within the healthcare system can all influence the accuracy of how these codes get assigned (1–3). There have been previous studies aiming to assess the validity of ICD codes for clinical research, but the results have been mainly inconclusive. Some studies attest to their use in research (26–33) while others argue they are not reliable (31,34–37). As new data sources become available and are combined with these clinically derived annotations, further investigation of the utility and validity of this coding is essential when exploring specific disease spaces.

One strategy to reduce biases and increase accuracy while evaluating ICD codes as proxies for diagnosis is grouping. ICD code grouping methods have been successful at categorizing patients into diseases where there’s a lack of coding consensus, and therefore, provide better insight into a patient’s medical condition, improving the generalizability of EHR-based research and the predictive power of clinical outcomes models (38–41). The pediatric complex chronic condition (CCC) v2 system is an established grouping methodology for identifying patients with CCCs. According to Feudtner et al., CCC encompasses any medical condition anticipated to last at least 12 months (unless death occurs), involving multiple organ systems or one severely affected organ system requiring specialized pediatric care and possibly tertiary care hospitalization (42). This system categorizes a broad range of ICD-9 and ICD-10 codes into ten groups (cardiovascular, neonatal, respiratory, neurologic/neuromuscular, renal/urologic, gastrointestinal, hematologic/immunologic, metabolic, other congenital or genetic defects, and malignancy), in which patients can belong to one or more groups. This system has been previously used in pediatric research, such as risk analysis and prediction of disease outcomes(42).

With large clinical datasets there is a need for methods that quantify the dependencies between thousands of features from multiple modalities in an explainable, portable, and accurate way. Bayesian networks are probabilistic graphical models that allow us to visualize and measure the probabilistic dependencies between features. They use directed acyclic graphs where the variables are represented by nodes, and their conditional dependencies are represented by edges. Bayesian networks are adept at handling a wide range of inquiries, from simple tasks like predicting outcomes based on a single variable to more complex ones involving multiple variables and their interplay (43–45). This makes them powerful tools for risk analysis and decision support in healthcare.

In this study, we are interested in understanding the risk factors contributing to pediatric sepsis. Sepsis is among the leading causes of pediatric death worldwide, and in 2017, it accounted for 2.9 million deaths in children under five years of age (46). Sepsis is a syndrome of life-threatening organ dysfunction that occurs when the body’s natural response to infection damages tissues. Unfortunately, disease development remains poorly understood due to the highly varying signs and symptoms that are nonspecific, making sepsis difficult to diagnose in an accurate and timely manner (46). Another challenge involves the absence of universally accepted definitions to classify pediatric patients with sepsis. Diagnosing and categorizing pediatric patients with severe sepsis depends on factors such as patient age and the presence of comorbidities. This categorization is further influenced by factors such as provider judgment and varying coding practices between healthcare systems (41,47).

The gold standard methodology for classifying patients in cohort studies and outcomes research is chart review. However, obtaining physician-validated datasets is burdensome, expensive, and time-consuming. In this study, we explore the use of ICD code groupings into clinically meaningful categories to increase our capability of identifying and measuring risk factors associated with sepsis severity. We use Bayesian networks to compare ICD categories to chart review and its associations with pediatric sepsis outcomes. A concern about using billing codes for research purposes is that they represent administrative information rather than biological or clinical reality. At face value, if the codes are applied in a consistent manner, then they represent useful information about the patients that can be used in cohort selection and outcomes research. Further, the categorization of these codes should serve to mitigate some of the biases that may be introduced administratively.

## Methods

### Setting and Study Population

We identified children aged 18 years and younger who had a culture-documented *E. coli* infection and were hospitalized at Primary Children’s Hospital (PCH) in Salt Lake City, UT, between 2012 and 2019. PCH is a standalone children’s hospital with 289 beds, serving as a pediatric community hospital for Salt Lake County, Utah, and the sole pediatric tertiary care center in the Intermountain West region. Demographic and clinical data were sourced from the Intermountain Health Care Enterprise Data Warehouse, with medical records manually reviewed to verify diagnoses and validate electronic data for all patients. Given the study’s timeframe and subjects’ ages, ICD-9 and ICD-10 codes were collected for these patients. The Institutional Review Boards of the University of Utah and Primary Children’s Hospitals approved this study with a waiver of informed consent as the data has been analyzed anonymously (IRB#00097637).

### Diagnosis Code categorization

1359 distinct ICD codes were collected by collating the patient’s records. Given the large number and the diversity of the coding terms, three different grouping strategies were implemented to reduce the number of terms by aggregating them based on clinical similarity.

### Data-Driven ICD code categorization: *sepsis-associated body systems*

Infection-associated codes were categorized into four body system groups (lower genitourinary tract, central nervous system, gastrointestinal, and blood) using expert knowledge. These body systems were selected because they represent the primary pathways through which severe infection typically arises. The infection-related diagnosis codes in each group were searched for within the list of patient diagnoses using a pattern-matching technique to aid collection of terms. For example, a list of all the central nervous system ICD codes relating to infection found within our cohort was created by searching all the patient diagnoses using prefixes, suffixes, and root words such as “cereb,” “brain,” “crani-,” etc to create a list for manual inspection. We constructed a presence-absence matrix where 1 represents a patient found to have any of the ICD codes in a given group, and 0 (an absence) if no ICD codes were found for that particular group. We repeated this process for each of the four groups.

### Data-Driven ICD code categorization: *Frequency of medical terms and context*

After compiling the terms directly associated with infection, we extracted the frequency of the remaining terms across all patients in our cohort. The frequencies were further explored to identify over-occurrence and common clinical phenotypes. We specifically focused on medical concepts related to prematurity, gastrostomy status, and thrombocytopenia, as they are known to impact severe sepsis outcomes. Using a pattern-matching technique, we searched for ICD-9 and ICD-10 codes corresponding to each medical concept within the pool of patient diagnoses. We incorporated these new groupings into our previous presence-absence matrix using the NLP approach mentioned above.

### Feudtner’s complex chronic condition v2 system

A presence-absence matrix for each CCC category was generated through SQL queries applied to patient data using standard code sets (42). This study only used the renal/urologic, gastrointestinal, hematologic/immunologic, malignancy, and other congenital or genetic defect categories as diagnostic code categories, resulting in five additional groups.

### Chart Reviewed Feature*s*

The electronic medical records and paper charts were retrospectively reviewed to obtain data regarding clinical presentation, extent of disease, infection risk factors, test results, and patient outcomes. Study data were collected and managed using REDCap electronic data capture tools hosted at PCH(48). The definition for each chart reviewed category is given below:

- **Severe sepsis:** Children with an invasive *E. coli* infection and evidence of tissue hypoperfusion or organ dysfunction due to the infection were categorized as having severe sepsis (49).
- **Bacteremia:** A positive blood culture for *E. coli*.
- **Meningitis:** 1) a positive cerebrospinal fluid (CSF) culture for *E. coli*, or 2) positive BioFire FilmArray Meningitis Encephalitis Panel result for *E. coli*, or 3) a positive blood culture for *E. coli* and evidence of infection in the CSF (WBC>5 cells/mm^3^), or 4) a positive blood culture for *E. coli* and a clinical diagnosis of meningitis as determined by the pediatric infectious disease physician.
- **Preterm infant:** Infant born prior to 37 weeks gestational age.
- **Preterm under 12 months:** Patient born prematurely, under 12 months of age at time of admission.
- **Urinary tract infection (UTI):** 1) positive urine culture (defined as growth greater than 10^5^ CFU/mL) *and* urinalysis with evidence of pyuria (^3^ 5 white blood cells or +leukocyte esterase) or +nitrites per American Academy of Pediatrics guidance (50).
- **Cancer:** Children admitted with an *E. coli* infection who were currently being treated for cancer.
- **Thrombocytopenia calculation:** Platelet count less than 150,000/uL of blood.
- **Gastrostomy tube:** Validation of the set of ICD derived cases using review of notes.

### Bayesian Network

A Bayesian network was built using the resulting ICD code categories, chart-reviewed diagnosis, lab values, and clinical outcomes as features. A Hill-Climbing greedy search algorithm was used to determine the network structure, employing the BDE score-based learning algorithm to search for the directed acyclic graph that maximized the network score. The learning process was bootstrapped 500 times. The R package bnlearn, version 4.9.1, was used to build the network and propagate the learned structure of the data to construct joint probabilities tables. The visual representation of our network was obtained using the visNetwork package from R, version 2.1.2.

### Risk Analysis

Relative risk (RR) and absolute risk (AR) are commonly used metrics to assess the association between an exposure and an outcome (43,44). RR quantifies the strength of association by comparing the risk of an outcome in an exposed group to the risk of an outcome in the non-exposed group. A RR above 1 indicates a higher risk of the outcome compared to the non-exposed group. On the other hand, AR measures the actual risk of an outcome. It is the ratio of the probability of an outcome in exposed individuals to the total number of individuals with the outcome.

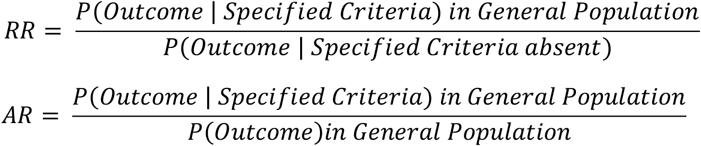

We calculated the RR and AR of a patient having severe sepsis, given the presence of one or more features by querying the Bayesian network. We repeated this process using bacteremia and meningitis as outcomes. The dataset was bootstrapped 1000 times to obtain the median RR, median AR, and 95% confidence intervals for each query. The risk calculations were performed using the gRain package in R, version 1.4.1.

## Results

### Patient Demographics

The overall patient population included in this study is described in Table 1. We identified 280 children with an *E. coli* infection treated at PCH over the 7-year study period. One-hundred and six cases (37.9%) occurred in children 5 years and older, and 85 (30.4) cases occurred in children 30 days to 2 years old (Table 1). In this study, age, ethnicity, and gender were not evaluated as risk factors. The most common clinical characteristic in our cohort was bacteremia (187 children; 66.8%), followed by severe sepsis (50 children; 17.9%) and meningitis (18 children; 6.4%: Table 1).

**Table 1.** Demographic and clinical characteristics of patients included in this study (n=280).

| Demographic Characteristics | Value (n = 280) |
| --- | --- |
| Age, median (IQR), months | 30.4 (1.8-134.8) |
| Age group, n (%), years: |  |
| Newborn | 51 (18.2%) |
| 30 days to 2 years | 85 (30.4%) |
| 2 years to 5 years | 38 (13.6%) |
| 5 years and older | 106 (37.9%) |
| Sex (n %): |  |
| Male | 132 (47.1%) |
| Female | 148 (52.9%) |
| Race, n (%): |  |
| American Indian or Alaska Native | 9 (3.2%) |
| Asian | 3 (1.1%) |
| Black or African American | 7 (2.5%) |
| Native Hawaiian or Other Pacific Islander | 12 (4.3%) |
| White | 236 (84.3%) |
| Other | 1 (0.4%) |
| Unknown/Declined | 12 (4.3%) |
| Ethnicity, n (%): |  |
| Hispanic, Latino, or Spanish Origin | 63 (22.5%) |
| Not Hispanic, Latino, or Spanish Origin | 210 (75%) |
| Unknown/Declined | 7 (2.5%) |
| Clinical Characteristics, n (%): |  |
| Severe Sepsis | 50 (17.9%) |
| Meningitis | 18 (6.4%) |
| Bacteremia | 187 (66.8%) |

### Bayesian Network

The final Bayesian network resulted in 21 features after removing those present in less than 5% or more than 95% of the cohort. Five nodes represent CCCs v2, seven nodes represent data-driven diagnoses, and nine nodes represent chart-reviewed diagnoses, including the severe sepsis outcome. Unlike traditional statistics, the Bayesian network allows us to visualize the relationships between all the variables as well as the association of the variables with the outcome (Figure 1). A univariable analysis was performed using the Bayesian network (Figure 2). This allowed for the comparison between the effects of data-generated variables versus chart-reviewed variables on an outcome. The RR of severe sepsis was increased more than two-fold by the presence of chart-review thrombocytopenia, data-driven thrombocytopenia, data-driven gastrointestinal infection, data-driven gastrostomy tube, and chart-reviewed gastrostomy tube. And the RRs of severe sepsis, given the presence of related data-driven and chart review diagnoses, are comparable. A web application was created to allow users to explore and query the Bayesian network. It was developed and deployed using the R package shiny, version 1.8.0, and is publicly accessible on a server at: https://lulu-valdez.shinyapps.io/sepsis_clinical_shinyapp/.

**Figure 1.**
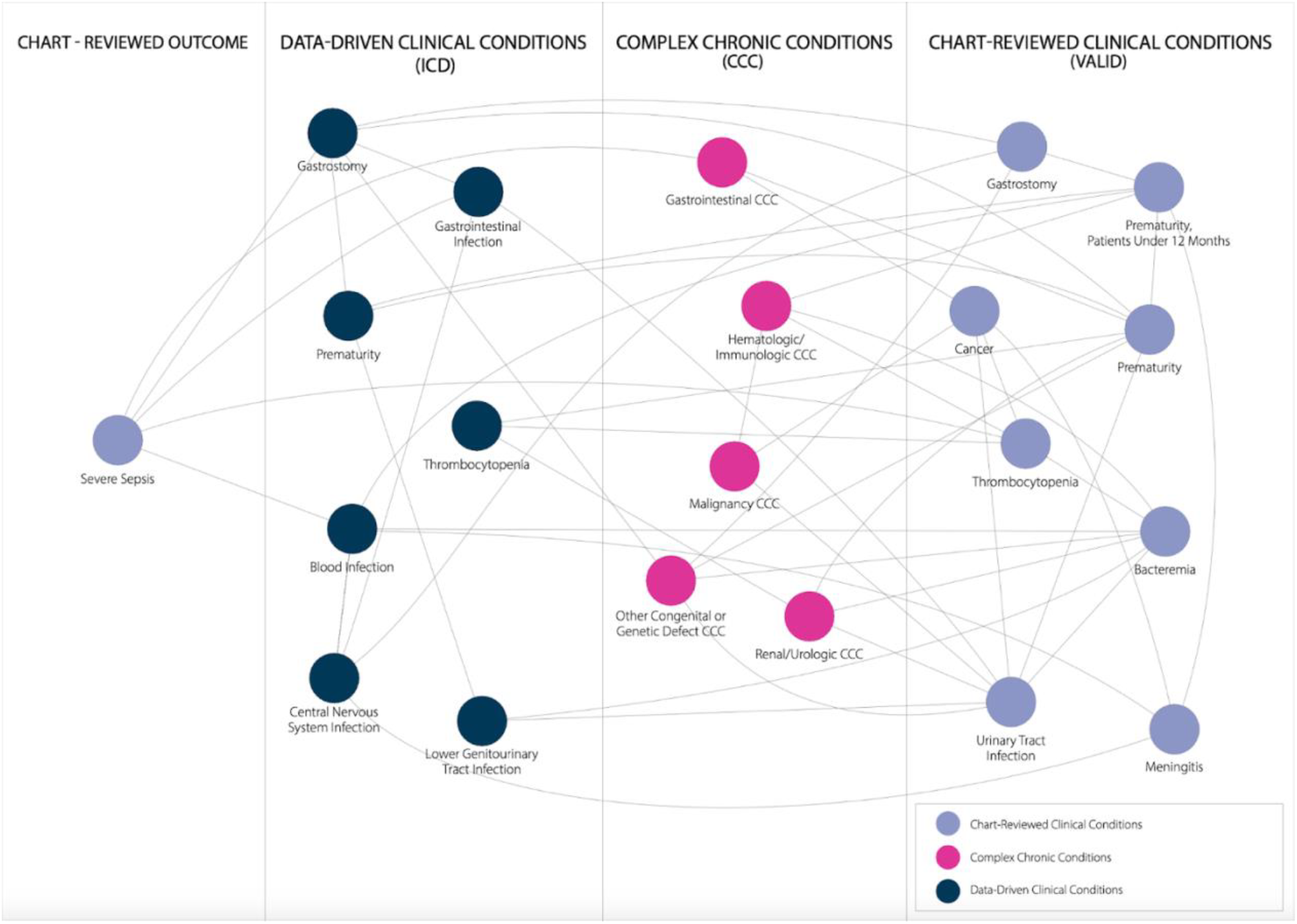
Bayesian network of clinical manifestations related to severe sepsis. Each node represents clinical categories (see methods), and lines represent the conditional dependencies between the nodes. Navy blue nodes represent data-driven clinical conditions, pink nodes represent Feudtner et al.’s CCC v2 system, and light purple nodes represent chart-reviewed clinical conditions.

**Figure 2.**
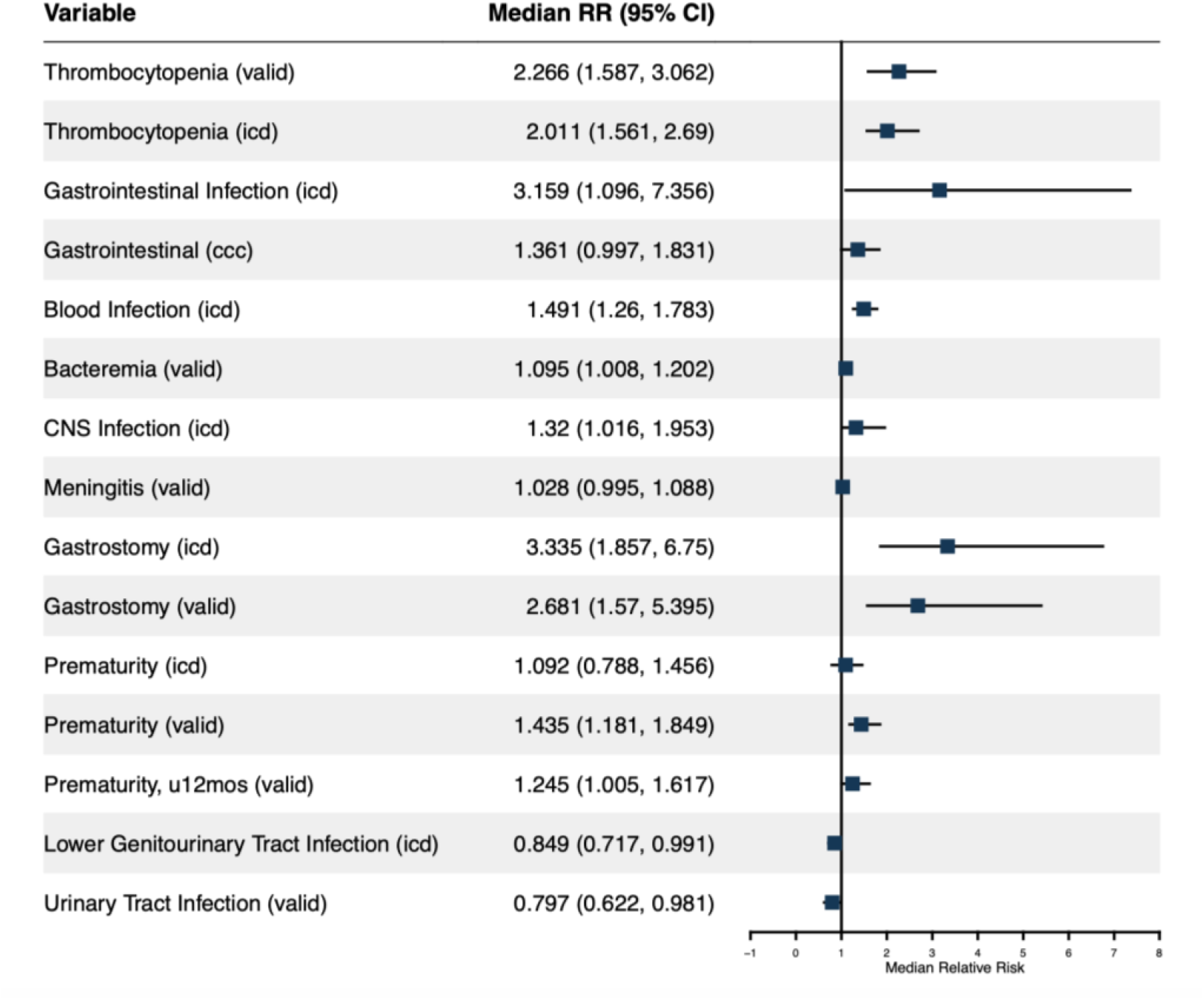
Relative risk of severe sepsis in pediatric patients. This figure shows a univariable analysis using the Bayesian network. Median relative risk scores and 95% confidence intervals were obtained after bootstrapping the network 1000 times.

### Sensitivity Analysis

The sensitivity analysis shown in Table 2 compares the difference between data-driven variables, the associated chart-reviewed variables, and their conditional dependency on severe sepsis. We analyzed nine pairs of nodes and identified five pairs of data-driven v. chart-review nodes as being significantly similar using Pearson’s Chi-squared test. The difference in the association between the two variables and severe sepsis was measured using the odds ratio (OR). OR = 1 indicates that the two predictors have an equal strength of association with the outcome. The closer the OR is to one, the more similar the two variables are to each other regarding their dependency on severe sepsis.

**Table 2.** Sensitivity analysis of data-driven variables, chart-review variables, and CCC variables.

| Variables | Outcome | Adjusted P-Value | Odds Ratio |
| --- | --- | --- | --- |
| Gastrointestinal Infection (icd)/<br>Gastrointestinal (ccc) | Severe Sepsis | <0.05 | 0.0991 |
| Prematurity (icd)/<br>Prematurity (valid) | Severe Sepsis | 1 | 0.5833 |
| Prematurity (icd)/<br>Prematurity, u12mos (valid) | Severe Sepsis | 1 | 0.7917 |
| Lower Genitourinary Tract<br>Infection (icd)/<br>Urinary Tract Infection (valid) | Severe Sepsis | 1 | 0.8863636 |
| Blood Infection (icd)/<br>Bacteremia (valid) | Severe Sepsis | 1 | 1.1282 |
| Prematurity(valid)/<br>Prematurity, u12mos (valid) | Severe Sepsis | 1 | 1.3571 |
| CNS Infection (icd)/<br>Meningitis (valid) | Severe Sepsis | 1 | 1.3623 |
| Gastrostomy (icd)/<br>Gastrostomy (valid) | Severe Sepsis | 1 | 2.4706 |
| Thrombocytopenia (valid)/<br>Thrombocytopenia (icd) | Severe Sepsis | <0.05 | 4.9916 |

This table represents a bivariate analysis performed using traditional statistical methods. Significant differences were assessed using Pearson's Chi-squared test. P-values were adjusted for multiple comparisons using the Bonferroni correction method. P-value < 0.05 indicates independence of association between each category and severe sepsis. Darker red represents decreasing OR while darker blue represents increasing OR.

## Discussion

We used the power of Bayesian networks to compare the effectiveness of data-driven ICD code categories as risk factors for sepsis-related outcomes and compared them to chart-reviewed diagnoses. Our Bayesian network identified four data-driven ICD code groupings that performed similarly to chart-reviewed diagnoses in predicting the risk of severe sepsis (Figure 1 and Table 2). The data-driven groupings were central nervous system infection, blood infection, lower genitourinary tract infection, and prematurity. Prematurity was compared to two different chart-reviewed prematurity variables (Prematurity and Prematurity, under 12 months), causing five pairs of variables to be identified but only four data-driven ICD code groupings.

We note that there are circumstances in which data-driven ICD groupings are less reliable than chart review. In our dataset, a grouping of ICD codes related to thrombocytopenia identified 37 cases, whereas chart review identified 89. This difference is seen when comparing the two categories with the outcomes. The most likely cause for this observation is that chart review used laboratory values to identify cases of thrombocytopenia, whereas the data-driven ICD code grouping only included the cases that were coded as thrombocytopenia as an indication for testing, but not those that were tested for other indications without a thrombocytopenia ICD code in their chart. We include the comparison of similar but non-overlapping groupings of terms to explore the impact on the outcomes using the Bayesian network. We created a grouping of terms involved in gastrointestinal infections and compared this grouping to the Gastrointestinal CCC category, which includes congenital anomalies, chronic liver disease/cirrhosis, and inflammatory bowel disease. While both of these groupings are connected in the network, it can be seen that they have very different impacts on the outcome. A gastrointestinal infection increases the risk of severe sepsis outcome threefold, but a chronic GI condition does not impact the outcome. This is to be expected.

In our exploration, we found a situation in which a temporal relationship might be missed by data-driven ICD groupings. ICD codes relating to prematurity were grouped in order to investigate infection relating to preterm infants. However, our cohort contained individuals from birth to 18 years of age, and therefore, our grouping included children who were premature at birth but who had the *E*.*coli* infection occur later in life. Chart review was capable of isolating the individuals who were infants when faced with infection with a higher level of certainty.

The use of data-driven ICD code groupings holds significant promise for understanding and predicting various medical outcomes. Bayesian networks offer a probabilistic framework that allows for the integration of diverse sources of data and the modeling of complex relationships between variables, thereby enhancing the accuracy and reliability of predictive models. Our investigation revealed key groupings that exhibited notable predictive capabilities in relation to sepsis-related outcomes. By leveraging Bayesian networks, we were able to effectively compare the performance of these data-driven groupings against chart-reviewed diagnoses, highlighting their potential utility in healthcare analytics.

## Conclusions

Our study demonstrates the potential of using data-driven ICD code groupings within Bayesian networks to effectively predict outcomes, particularly severe sepsis. We identified four distinct groupings—central nervous system infection, blood infection, lower genitourinary tract infection, and prematurity—that showed comparable performance to chart-reviewed diagnoses. However, it is important to recognize the limitations of solely relying on data-driven methodologies. There are instances where chart review may offer a more comprehensive and accurate assessment, particularly when dealing with temporal clinical findings or when the use of laboratory values is warranted. Thus, while data-driven approaches have proven valuable for outcome prediction within the context of Bayesian networks, certain situations require they be complemented by traditional methods to ensure robustness and reliability. Using this combination of this data-driven classification of patients by ICD code with Bayesian networks to explore the conditional probability topology of data enables rapid exploration of clinical datasets. This process can be iterative and amenable to the integration of further datasets.

## Data Availability

All data produced in the present study are available upon reasonable request to the authors.

https://lulu-valdez.shinyapps.io/sepsis_clinical_shinyapp/

## Acknowledgments

The authors would like to thank Juliana Klein for her assistance in designing the Bayesian network figure presented in this manuscript. LV is supported by the NLM Training grant T15LM007124 awarded to KE. This study is also funded by the NIH R01 GM134331 grant and Department of Defense award W81XWH-22-1-0800 (SC210103) awarded to MAM.

## Notes

### Competing Interest Statement

The authors have declared no competing interest.

### Funding Statement

Lourdes Valdez is supported by the NLM Training grant T15LM007124 awarded to Karen Eilbeck. The study was also funded by the NIH R01 GM134331 grant and Department of Defense award W81XWH-22-1-0800 (SC210103) awarded to Matthew A. Mulvey

### Author Declarations

The Institutional Review Boards of the University of Utah and Primary Childrens Hospitals approved this study with a waiver of informed consent as the data has been analyzed anonymously (IRB#00097637).

## References

1. Sauer CM, Chen LC, Hyland SL, Girbes A, Elbers P, Celi LA. Leveraging electronic health records for data science: common pitfalls and how to avoid them. Lancet Digit Health. 2022 Dec;4(12):e893–8.

2. Li HC, Chen YW, Chen TJ, Chiou SH, Hwang SJ. The role of patient records in research: A bibliometric analysis of publications from an academic medical center in Taiwan. J Chin Med Assoc. 2021 Jul 1;84(7):718–21.

3. Johnson EK, Nelson CP. Values and pitfalls of the use of administrative databases for outcomes assessment. J Urol. 2013 Jul;190(1):17–8.

4. Goto T, Camargo CA Jr, Faridi MK, Freishtat RJ, Hasegawa K. Machine Learning-Based Prediction of Clinical Outcomes for Children During Emergency Department Triage. JAMA Netw Open. 2019 Jan 4;2(1):e186937.

5. Zhang S, Yang F, Wang L, Si S, Zhang J, Xue F. Personalized prediction for multiple chronic diseases by developing the multi-task Cox learning model. PLoS Comput Biol. 2023 Sep;19(9):e1011396.

6. Witmer CM, Huang YS, Lynch K, Raffini LJ, Shah SS. Off-label recombinant factor VIIa use and thrombosis in children: a multi-center cohort study. J Pediatr. 2011 May;158(5):820–5.e1.

7. Weiss AK, Hall M, Lee GE, Kronman MP, Sheffler-Collins S, Shah SS. Adjunct corticosteroids in children hospitalized with community-acquired pneumonia. Pediatrics. 2011 Feb;127(2):e255–63.

8. Srivastava R, Berry JG, Hall M, Downey EC, O’Gorman M, Dean JM, et al. Reflux related hospital admissions after fundoplication in children with neurological impairment: retrospective cohort study. BMJ. 2009 Nov 18;339:b4411.

9. Matlow AG, Baker GR, Flintoft V, Cochrane D, Coffey M, Cohen E, et al. Adverse events among children in Canadian hospitals: the Canadian Paediatric Adverse Events Study. CMAJ. 2012 Sep 18;184(13):E709–18.

10. Martin ET, Kuypers J, Wald A, Englund JA. Multiple versus single virus respiratory infections: viral load and clinical disease severity in hospitalized children. Influenza Other Respi Viruses. 2012 Jan;6(1):71–7.

11. Shah SS, Hall M, Newland JG, Brogan TV, Farris RWD, Williams DJ, et al. Comparative effectiveness of pleural drainage procedures for the treatment of complicated pneumonia in childhood. J Hosp Med. 2011 May;6(5):256–63.

12. Shah SS, Hall M, Srivastava R, Subramony A, Levin JE. Intravenous immunoglobulin in children with streptococcal toxic shock syndrome. Clin Infect Dis. 2009 Nov 1;49(9):1369–76.

13. Simon TD, Hall M, Michael Dean J, Kestle JRW, Riva-Cambrin J. Reinfection following initial cerebrospinal fluid shunt infection: Clinical article. J Neurosurg Pediatr. 2010 Sep 1;6(3):277–85.

14. Simon TD, Berry J, Feudtner C, Stone BL, Sheng X, Bratton SL, et al. Children with complex chronic conditions in inpatient hospital settings in the United States. Pediatrics. 2010 Oct;126(4):647–55.

15. Setty BA, O’Brien SH, Kerlin BA. Pediatric venous thromboembolism in the United States: a tertiary care complication of chronic diseases. Pediatr Blood Cancer. 2012 Aug;59(2):258–64.

16. Edwards JD, Houtrow AJ, Vasilevskis EE, Rehm RS, Markovitz BP, Graham RJ, et al. Chronic conditions among children admitted to U.S. pediatric intensive care units: their prevalence and impact on risk for mortality and prolonged length of stay*. Crit Care Med. 2012 Jul;40(7):2196–203.

17. Shah SS, Ten Have TR, Metlay JP. Costs of treating children with complicated pneumonia: a comparison of primary video-assisted thoracoscopic surgery and chest tube placement. Pediatr Pulmonol. 2010 Jan;45(1):71– 7.

18. Coffin SE, Leckerman K, Keren R, Hall M, Localio R, Zaoutis TE. Oseltamivir shortens hospital stays of critically ill children hospitalized with seasonal influenza: a retrospective cohort study. Pediatr Infect Dis J. 2011 Nov;30(11):962–6.

19. Berry JG, Hall DE, Kuo DZ, Cohen E, Agrawal R, Feudtner C, et al. Hospital utilization and characteristics of patients experiencing recurrent readmissions within children’s hospitals. JAMA. 2011 Feb 16;305(7):682–90.

20. Advani S, Reich NG, Sengupta A, Gosey L, Milstone AM. Central line-associated bloodstream infection in hospitalized children with peripherally inserted central venous catheters: extending risk analyses outside the intensive care unit. Clin Infect Dis. 2011 May;52(9):1108–15.

21. Benneyworth BD, Gebremariam A, Clark SJ, Shanley TP, Davis MM. Inpatient health care utilization for children dependent on long-term mechanical ventilation. Pediatrics. 2011 Jun;127(6):e1533–41.

22. Khan A, Uddin S, Srinivasan U. Chronic disease prediction using administrative data and graph theory: The case of type 2 diabetes. Expert Syst Appl. 2019 Dec 1;136:230–41.

23. Lu H, Uddin S. Disease Prediction Using Graph Machine Learning Based on Electronic Health Data: A Review of Approaches and Trends. Healthcare (Basel) [Internet]. 2023 Apr 4;11(7). Available from: 10.3390/healthcare11071031

24. Lu H, Uddin S. A weighted patient network-based framework for predicting chronic diseases using graph neural networks. Sci Rep. 2021 Nov 19;11(1):22607.

25. Kurbasic I, Pandza H, Masic I, Huseinagic S, Tandir S, Alicajic F, et al. The advantages and limitations of international classification of diseases, injuries and causes of death from aspect of existing health care system of bosnia and herzegovina. Acta Inform Med. 2008;16(3):159–61.

26. Jolley RJ, Sawka KJ, Yergens DW, Quan H, Jetté N, Doig CJ. Validity of administrative data in recording sepsis: a systematic review. Crit Care. 2015 Apr 6;19(1):139.

27. Liu B, Hadzi-Tosev M, Liu Y, Lucier KJ, Garg A, Li S, et al. Accuracy of International Classification of Diseases, 10th Revision Codes for Identifying Sepsis: A Systematic Review and Meta-Analysis. Crit Care Explor. 2022 Nov;4(11):e0788.

28. Tavakoli H, Chen W, Sin DD, FitzGerald JM, Sadatsafavi M. Predicting Severe Chronic Obstructive Pulmonary Disease Exacerbations. Developing a Population Surveillance Approach with Administrative Data. Ann Am Thorac Soc. 2020 Sep;17(9):1069–76.

29. McGrew KM, Homco JB, Garwe T, Dao HD, Williams MB, Drevets DA, et al. Validity of International Classification of Diseases codes in identifying illicit drug use target conditions using medical record data as a reference standard: A systematic review. Drug Alcohol Depend. 2020 Mar 1;208:107825.

30. Peng M, Eastwood C, Boxill A, Jolley RJ, Rutherford L, Carlson K, et al. Coding reliability and agreement of International Classification of Disease, 10th revision (ICD-10) codes in emergency department data. Int J Popul Data Sci. 2018 Jul 26;3(1):445.

31. Higgins TL, Deshpande A, Zilberberg MD, Lindenauer PK, Imrey PB, Yu PC, et al. Assessment of the Accuracy of Using ICD-9 Diagnosis Codes to Identify Pneumonia Etiology in Patients Hospitalized With Pneumonia. JAMA Netw Open. 2020 Jul 1;3(7):e207750.

32. Park J, Kwon S, Choi EK, Choi YJ, Lee E, Choe W, et al. Validation of diagnostic codes of major clinical outcomes in a National Health Insurance database. J Interv Card Electrophysiol. 2019 Nov 20;20(1):1–7.

33. Callahan K, Acharya Y, Hollenbeak CS. Validity of ICD codes to identify do-not-resuscitate orders among older adults with heart failure: A single center study. PLoS One. 2023 Mar 13;18(3):e0283045.

34. Rhee C, Murphy MV, Li L, Platt R, Klompas M, Centers for Disease Control and Prevention Epicenters Program. Comparison of trends in sepsis incidence and coding using administrative claims versus objective clinical data. Clin Infect Dis. 2015 Jan 1;60(1):88–95.

35. Endrich O, Triep K, Schlapbach LJ, Posfay-Barbe KM, Heininger U, Giannoni E, et al. Sensitivity of ICD coding for sepsis in children-a population-based study. Intensive Care Med Paediatr Neonatal. 2023 Jun 13;1(1):5.

36. Kelly GF, Makhoul T, Zammit CG, Jones CMC, Acquisto NM. The accuracy of ICD-CM codes to identify thromboembolic events for clinical outcomes research. J Am Coll Clin Pharm. 2021 Jan;4(1):40–6.

37. Beam KS, Lee M, Hirst K, Beam A, Parad RB. Specificity of International Classification of Diseases codes for bronchopulmonary dysplasia: an investigation using electronic health record data and a large insurance database. J Perinatol. 2021 Apr;41(4):764–71.

38. Hagström H, Adams LA, Allen AM, Byrne CD, Chang Y, Grønbaek H, et al. Administrative Coding in Electronic Health Care Record-Based Research of NAFLD: An Expert Panel Consensus Statement. Hepatology. 2021 Jul;74(1):474–82.

39. Zhang L, Zhang Y, Cai T, Ahuja Y, He Z, Ho YL, et al. Automated grouping of medical codes via multiview banded spectral clustering. J Biomed Inform. 2019 Dec;100:103322.

40. Kansal A, Gao M, Balu S, Nichols M, Corey K, Kashyap S, et al. Impact of diagnosis code grouping method on clinical prediction model performance: A multi-site retrospective observational study. Int J Med Inform. 2021 Jul;151:104466.

41. Magill SS, Sapiano MRP, Gokhale R, Nadle J, Johnston H, Brousseau G, et al. Epidemiology of Sepsis in US Children and Young Adults. Open Forum Infect Dis. 2023 May;10(5):ofad218.

42. Feudtner C, Feinstein JA, Zhong W, Hall M, Dai D. Pediatric complex chronic conditions classification system version 2: updated for ICD-10 and complex medical technology dependence and transplantation. BMC Pediatr. 2014 Aug 8;14:199.

43. Zimmerman RM, Hernandez EJ, Watkins WS, Blue N, Tristani-Firouzi M, Yandell M, et al. An Explainable Artificial Intelligence Approach for Discovering Social Determinants of Health and Risk Interactions for Stroke in Patients With Atrial Fibrillation. Am J Cardiol. 2023 Aug 15;201:224–6.

44. Kiser AC, Schliep KC, Hernandez EJ, Peterson CM, Yandell M, Eilbeck K. An artificial intelligence approach for investigating multifactorial pain-related features of endometriosis. PLoS One. 2024 Feb 21;19(2):e0297998.

45. Wesołowski S, Lemmon G, Hernandez EJ, Henrie A, Miller TA, Weyhrauch D, et al. An explainable artificial intelligence approach for predicting cardiovascular outcomes using electronic health records. PLOS Digit Health [Internet]. 2022 Jan 18;1(1). Available from: 10.1371/journal.pdig.0000004

46. Global report on the epidemiology and burden of sepsis: current evidence, identifying gaps and future directions. Geneva: World Health Organization; 2020.

47. Rudd KE, Kissoon N, Limmathurotsakul D, Bory S, Mutahunga B, Seymour CW, et al. The global burden of sepsis: barriers and potential solutions. Crit Care. 2018 Sep 23;22(1):232.

48. Harris PA, Taylor R, Minor BL, Elliott V, Fernandez M, O’Neal L, et al. The REDCap consortium: Building an international community of software platform partners. J Biomed Inform. 2019 Jul;95:103208.

49. Dellinger RP, Levy MM, Rhodes A, Annane D, Gerlach H, Opal SM, et al. Surviving sepsis campaign: international guidelines for management of severe sepsis and septic shock: 2012. Crit Care Med. 2013 Feb;41(2):580–637.

50. Subcommittee on Urinary Tract Infection, Steering Committee on Quality Improvement and Management, Roberts KB. Urinary tract infection: clinical practice guideline for the diagnosis and management of the initial UTI in febrile infants and children 2 to 24 months. Pediatrics. 2011 Sep;128(3):595–610.

